# The impact of extranodal spread on overall and recurrence-free survival in patients with clinically early-stage cervical cancer, but with tumor positive pelvic lymph nodes after radical hysterectomy

**DOI:** 10.64898/2026.09.02.26362042

**Authors:** Roos Gerarda Francina Maria van der Ven, Hans Hartmuth Bernhard Wenzel, Maaike Anne van der Aa, Constantijne Helene Mom, Nikki Bo Thuijs, Jacobus van der Velden

## Abstract

**Objective:** Extranodal spread is an adverse prognostic factor in several cancers. In clinically early-stage cervical cancer and positive lymph nodes after radical hysterectomy, extranodal spread is routinely registered in pathology reports, but it does not influence adjuvant treatment decisions. This nationwide study assessed the prognostic value of extranodal spread.

**Methods:** Patients diagnosed between 2009 and 2024 with clinical early-stage cervical cancer (TNM T1 to T2a2) and pathologically positive regional lymph nodes after radical hysterectomy were identified from the Netherlands Cancer Registry. Cox proportional-hazard models assessed differences in overall survival and recurrence-free survival. A Bayesian Cox model with informative prior was applied to complement overall survival analysis. Recurrence patterns were compared by extranodal spread status.

**Results:** Among 408 patients, 46 (11.3%) had extranodal spread. In multivariable frequentist analysis, extranodal spread was significantly associated with poorer recurrence-free survival (hazard ratio [HR] 1.97, 95% confidence interval [CI] 1.11–3.49), but not with overall survival (HR 1.37, CI 0.78–2.41). Bayesian analysis showed a significant difference in overall survival (HR 1.63, CI 1.14–2.27). Among patients with recurrence, the distribution of locoregional and distant recurrence patterns did not significantly differ between those with and without extranodal spread (p=0.09).

**Conclusion:** Extranodal spread is associated with poorer recurrence-free survival in clinically early-stage cervical cancer treated with radical hysterectomy. For overall survival, results were directionally consistent but dependent on the analytical approach, likely reflecting limited statistical power. These findings suggest a role for extranodal spread in risk stratification and follow-up, warranting further investigation before informing treatment decisions.

**Disclosures:** The authors have no conflicts of interest to disclose.

**Key messages:** *What is already known on this topic:* Extranodal spread is an established adverse prognostic factor in several malignancies. In clinically early-stage cervical cancer treated with radical hysterectomy, extranodal spread is routinely reported when lymph nodes are positive, although it does not currently influence adjuvant treatment decisions.

*What this study adds:* In this nationwide cohort of patients with clinically early-stage cervical cancer treated with radical hysterectomy, extranodal spread was associated with poorer recurrence-free survival. For overall survival, estimates consistently suggested worse outcomes in patients with extranodal spread, although statistical significance depended on the analytical approach.

*How this study might affect research, practice or policy:* These findings suggest that extranodal spread may be a clinically relevant prognostic marker in clinically early-stage cervical cancer and could contribute to improved risk stratification, follow-up strategies, and future evaluation of adjuvant treatment guidelines.

## Introduction

Cervical cancer constitutes a significant global health burden, ranking as the fourth most common cancer worldwide, causing around 570,000 new cases and 311,000 deaths each year.^1^ In patients with clinically early-stage disease (T1 to T2a2) as determined by gynecological examination and imaging, radical hysterectomy is primarily recommended.^2^ The presence of positive pelvic lymph nodes following radical hysterectomy is a well-established adverse prognostic factor.^3^ Other nodal parameters, including the number of involved nodes, lymph node ratio, size of metastases, and extranodal spread, have also been explored for their potential prognostic relevance in clinically early-stage cervical cancer.^4-6^ In several other cancers, including head and neck cancer, vulvar cancer, and ovarian cancer, extranodal spread is already an established adverse prognostic factor.^7-9^ Consequently, in vulvar and head-and-neck cancers, extranodal spread is systematically reported, as its presence directly influences and guides tailored adjuvant treatment strategies.^10, 11^

For cervical cancer, the European Society of Gynaecological Oncology (ESGO) recommends systematic reporting of nodal characteristics for quality assurance purposes.^2^ However, with the exception of positive common iliac nodes, such parameters currently do not have any impact on the type of adjuvant therapy or follow-up.^2^ This may reflect that their potential prognostic significance remains uncertain. Some studies have reported no survival difference between patients with and without extranodal spread,^12, 13^ whereas others have demonstrated significantly worse survival among patients with extranodal spread compared to those with intranodal metastases only.^4, 14, 15^ Many of these studies, however, have limitations, such as highly selected patient populations, small cohorts, or reliance on historical, single-center databases. Moreover, they did not systematically assess patterns of recurrence, which could be particularly relevant, if extranodal spread status was ever to be incorporated into adjuvant treatment strategies after radical hysterectomy.

The currently limited use of extranodal spread to guide postoperative adjuvant treatment and follow-up raises questions about the clinical relevance of this parameter, as well as about the evidence base supporting its systematic assessment and reporting. If extranodal spread proves to be a robust prognostic factor, it could support more personalized follow-up strategies after radical hysterectomy. With ESGO recommending structured documentation of extranodal spread in pathology reports, this parameter is apparently considered an important prognostic feature.^2^ Confirming its association with overall survival, recurrence-free survival, and recurrence patterns could therefore directly inform clinical practice and support more personalized adjuvant treatment decisions. This study adopts a nationwide perspective using real-world, population-based data to evaluate the prognostic value of extranodal spread in patients with clinically early-stage cervical cancer treated with radical hysterectomy and pathological lymph node assessment. Additionally, it aims to provide insights into recurrence patterns. By doing so, the study assesses the prognostic role of extranodal spread in cervical cancer.

## Methods

### Data collection

For this retrospective nationwide population-based cohort study, data from the Netherlands Cancer Registry (NCR) and the Dutch nationwide registry of histo- and cytopathology (PALGA) were used. The NCR is a population-based registry that includes data on all newly diagnosed malignancies in the Netherlands, since 1989.^16^ Data managers follow comprehensive reporting manuals and are specially trained to extract patient, tumor, and treatment information from medical records. Regular data validation procedures are conducted to ensure data quality. PALGA is the nationwide archive covering all pathology laboratories in the Netherlands.^17^ Excerpts of all histopathology and cytopathology reports are generated at the laboratories and transferred to the central database, where systematic quality and completeness checks are performed.

Data on patient and disease-related characteristics were collected, including year of diagnosis, age at time of diagnosis, tumor stage, histology, tumor differentiation grade, lympho-vascular space invasion, depth of invasion, tumor size, parametrial invasion, the number of lymph nodes examined, the number of positive lymph nodes, extranodal spread status, and type of adjuvant therapy (chemotherapy and/or radiotherapy). The anatomical site of the primary tumor and metastases is classified according to the 3rd edition of the International Classification of Diseases for Oncology (ICD-O).^18^ Tumor stage was determined using the Union for International Cancer Control (UICC) tumor-node-metastasis (TNM) classification, corresponding to the edition valid at the time of diagnosis.^19^ Overall survival is defined as the interval in months between diagnosis and death. Recurrence-free survival is defined as the length of time from date of hysterectomy to first locoregional recurrence or distant metastases or death due to any cause, whichever occurs first. Information on recurrence was limited to the first recorded recurrence event; subsequent recurrences were not available. Site of first recurrence was classified as locoregional or distant. Locoregional recurrence included recurrence at or near the primary tumor site and metastases to lymph nodes classified as regional according to the TNM system. Distant recurrence comprised distant metastases. In cases with multiple recurrence sites, classification was based on the most advanced manifestation, with distant recurrence taking precedence over locoregional recurrence. Follow-up on vital status was available up to January 2025, based on records from the Dutch Municipal Personal Records Database.

### Patient selection

All Dutch patients (≥18 years) diagnosed between January 2009 and December 2024 with clinical stage T1 through T2a cervical cancer, and without distant metastasis, of squamous cell carcinoma, adenocarcinoma or adenosquamous carcinoma histology, who underwent radical hysterectomy and pathological lymph node assessment as primary treatment and had at least one positive regional node, were included. Patients were excluded if they had received neoadjuvant therapy, as this may alter nodal status and extranodal spread.

### Analysis

Patient and disease-related characteristics are described using medians with interquartile ranges (IQR) for non-normally distributed continuous variables, and frequencies with percentages for categorical variables. Pearson’s chi-squared tests, Fisher’s exact test (for discrete variables with <5 observations) or Mann–Whitney U-tests were used to detect baseline differences in patient, tumor, and treatment characteristics.

Unadjusted Kaplan-Meier survival curves were generated to depict overall survival during the first five years after primary diagnosis and recurrence-free survival during the first five years post hysterectomy. Frequentist multivariable Cox proportional-hazard models were applied to analyze the impact of extranodal spread on overall survival and recurrence-free survival. Overall survival was analyzed in the entire cohort (patients diagnosed between 2009 and 2024), with vital status data available for 378 of 408 patients (92.6%). Recurrence-free survival could only be assessed in patients diagnosed between 2009 and 2019, the period for which recurrence data were collected, with recurrence data available for 265 of 305 patients (86.9%). Results are presented in (HR) with 95% confidence interval (CI).

Previous studies have shown that extranodal spread is relatively uncommon, resulting in a limited number of patients with extranodal spread available for analysis.^4, 12-15^ Consequently, effect estimates derived solely from the current dataset were expected to be associated with considerable uncertainty. As previously published literature on the association between extranodal spread and overall survival was available, a complementary Bayesian survival analysis was performed. Bayesian methods provide a principled framework for combining prior evidence with newly observed data and can be particularly informative when sample sizes are limited.^20^ The Bayesian analysis was therefore used as a complementary inferential approach to evaluate the consistency of the current findings in the context of existing evidence.

Informative prior distributions were derived from Horn et al., 2008,^4^ the only study reporting HR estimates for the association between extranodal spread on overall survival. The prior mean corresponded to the reported HR, transformed to the log scale, and the prior variance was calculated from the published 95%CI. All other covariates were assigned weakly informative normal priors centered at zero. Posterior inference was performed using Markov chain Monte Carlo sampling implemented in Stan using the *rstan* package. Four chains were run with 2,000 iterations each, including a burn-in of 500 iterations. Convergence was assessed using standard diagnostics. Bayesian analyses were restricted to overall survival, as no published estimates reporting HRs with corresponding CIs were available to inform an evidence-based prior for the association between extranodal spread and recurrence-free survival. Results are presented as posterior mean HR with 95% credibility intervals and posterior probabilities.

Selection of covariates for both frequentist and Bayesian Cox proportional-hazard models was guided by clinical relevance, considering their potential confounding effect and considering the number of parameters to be added in relation to the number of events.^21^ Patients are censored at either January 2025, the date of last visit or the date of emigration, whichever occurred first.

For patients who experienced a recurrence within five years, the site of first recurrence was recorded and categorized as locoregional or distant. The proportions of patients with any recurrence, locoregional recurrence, and distant recurrence were compared between patients with and without extranodal spread using chi-square tests.

Because in the Netherlands Cancer Registry capsular status was available for only part of the cohort (62.8%), additional data were extracted from free-text pathology reports by the first author. In 82 patients (20.1%), capsular status was not explicitly reported (appendix A.1). In these cases, absence of extranodal spread was assumed. A sensitivity analysis was performed comparing these patients with those with pathologically confirmed absence of extranodal spread.

All frequentist analyses were performed using Stata/SE version 17.0 (StataCorp LLC, College Station, TX, USA). Bayesian analyses were conducted in R version 4.5.1 (R Foundation for Statistical Computing, Vienna, Austria). Statistical tests were considered statistically significant at p < 0.05.

This study was approved by the Privacy Review Board of the Netherlands Cancer Registry (24/02/2025: 24-00647) and the Privacy Review Board of the Dutch Nationwide Network and Registry of Histo- and Cytopathology (PALGA) (29/01/2025: 2025-6). The data that support this study’s findings are available from the Netherlands Comprehensive Cancer Organisation (IKNL) and the Dutch nationwide registry of histo- and cytopathology (PALGA). Data from IKNL are used under license for this study and therefore not publicly available. (Analytical) code(s) are available upon (reasonable) request in consultation with the corresponding author. Data from PALGA are not publicly available due to PALGA regulations and patient privacy but may be accessed upon reasonable request and approval by PALGA. This research did not receive any specific grant from funding agencies in the public, commercial, or not-for-profit sectors.

## Results

A total of 408 patients with clinically early-stage cervical cancer and one or more positive regional lymph nodes after radical hysterectomy and pathological lymph node assessment were included in this study, of whom 46 (11.3%) had pathologically confirmed extranodal spread. Tumors with extranodal spread had a higher prevalence of parametrial invasion (28.3% vs. 12.4%; p=0.01) compared to those without extranodal spread. Moreover, patients with extranodal spread had a higher number of positive lymph nodes (median 2 vs. 1; p=0.01). The type of adjuvant treatment did not differ between the two groups (Table 1).

**Table 1.**
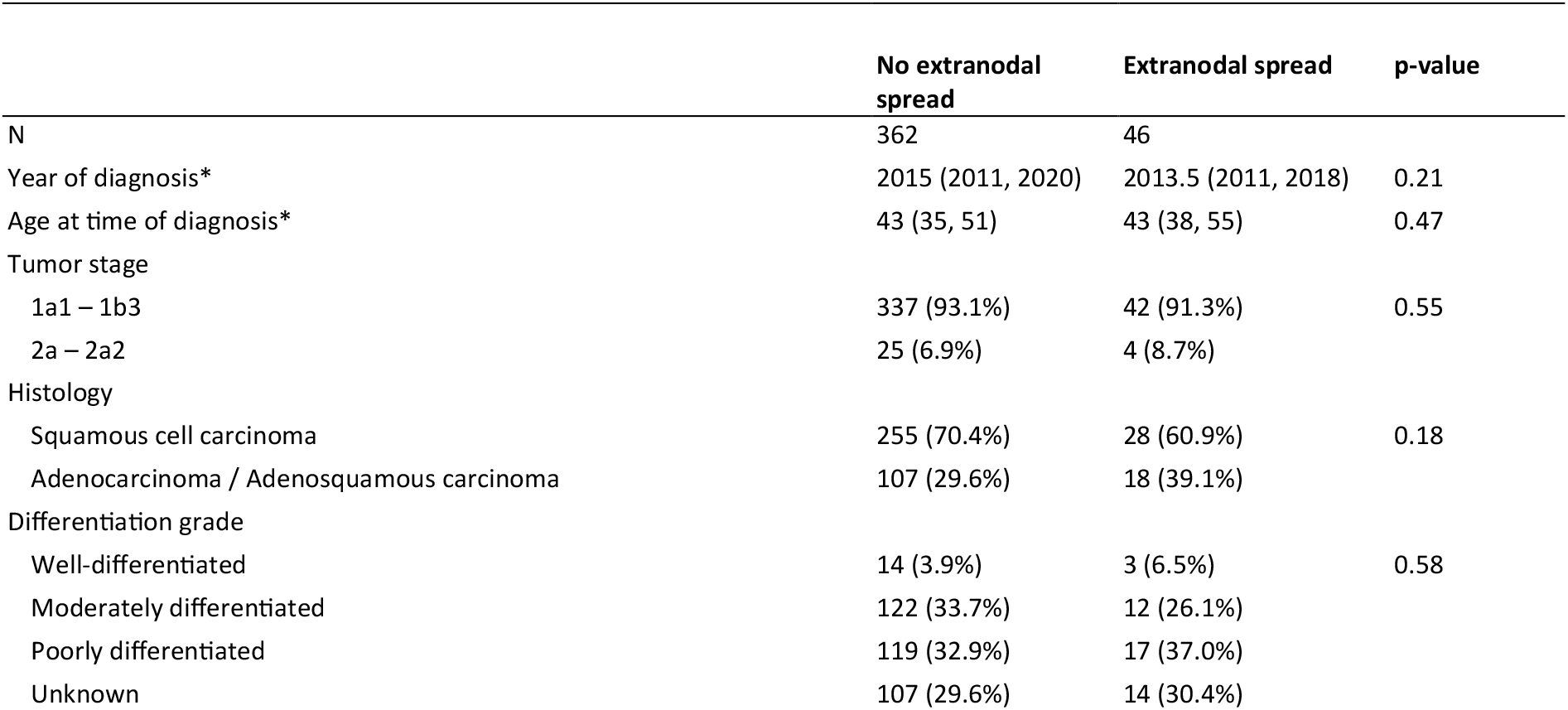
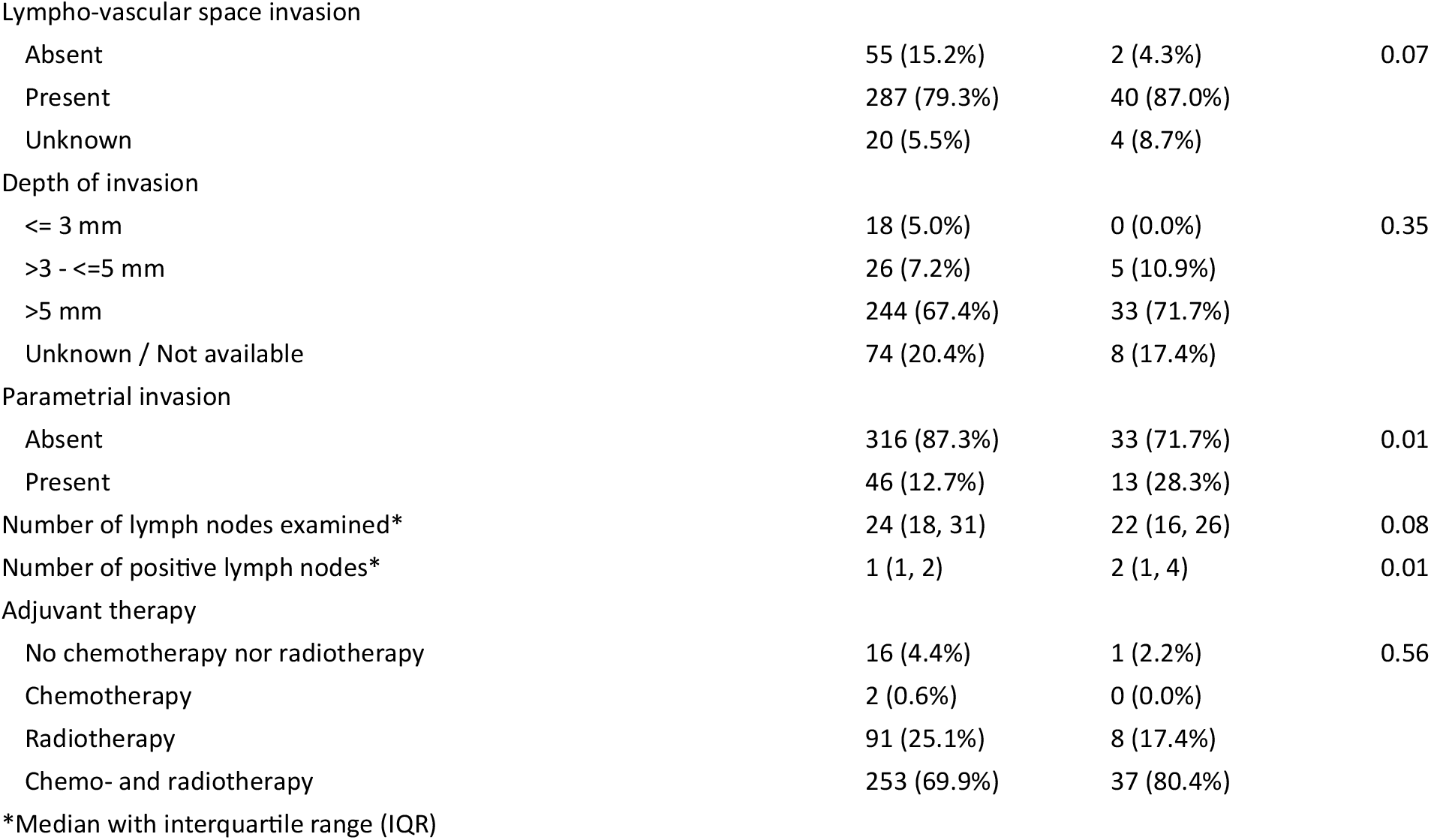
Patient and disease-related characteristics.

|  | No extranodal spread | Extranodal spread | p-value |
| --- | --- | --- | --- |
| N | 362 | 46 |  |
| Year of diagnosis* | 2015 (2011, 2020) | 2013.5 (2011, 2018) | 0.21 |
| Age at time of diagnosis* | 43 (35, 51) | 43 (38, 55) | 0.47 |
| Tumor stage |  |  |  |
| 1a1 – 1b3 | 337 (93.1%) | 42 (91.3%) | 0.55 |
| 2a – 2a2 | 25 (6.9%) | 4 (8.7%) |  |
| Histology |  |  |  |
| Squamous cell carcinoma | 255 (70.4%) | 28 (60.9%) | 0.18 |
| Adenocarcinoma / Adenosquamous carcinoma | 107 (29.6%) | 18 (39.1%) |  |
| Differentiation grade |  |  |  |
| Well-differentiated | 14 (3.9%) | 3 (6.5%) | 0.58 |
| Moderately differentiated | 122 (33.7%) | 12 (26.1%) |  |
| Poorly differentiated | 119 (32.9%) | 17 (37.0%) |  |
| Unknown | 107 (29.6%) | 14 (30.4%) |  |
| Lympho-vascular space invasion |  |  |  |
| Absent | 55 (15.2%) | 2 (4.3%) | 0.07 |
| Present | 287 (79.3%) | 40 (87.0%) |  |
| Unknown | 20 (5.5%) | 4 (8.7%) |  |
| Depth of invasion |  |  |  |
| <= 3 mm | 18 (5.0%) | 0 (0.0%) | 0.35 |
| >3 - <=5 mm | 26 (7.2%) | 5 (10.9%) |  |
| >5 mm | 244 (67.4%) | 33 (71.7%) |  |
| Unknown / Not available | 74 (20.4%) | 8 (17.4%) |  |
| Parametrial invasion |  |  |  |
| Absent | 316 (87.3%) | 33 (71.7%) | 0.01 |
| Present | 46 (12.7%) | 13 (28.3%) |  |
| Number of lymph nodes examined* | 24 (18, 31) | 22 (16, 26) | 0.08 |
| Number of positive lymph nodes* | 1 (1, 2) | 2 (1, 4) | 0.01 |
| Adjuvant therapy |  |  |  |
| No chemotherapy nor radiotherapy | 16 (4.4%) | 1 (2.2%) | 0.56 |
| Chemotherapy | 2 (0.6%) | 0 (0.0%) |  |
| Radiotherapy | 91 (25.1%) | 8 (17.4%) |  |
| Chemo- and radiotherapy | 253 (69.9%) | 37 (80.4%) |  |
| *Median with interquartile range (IQR) |  |  |  |

\*Median with interquartile range (IQR)

In the unadjusted survival analysis, no difference in overall survival was observed between patients with or without extranodal spread (p = 0.09). For patients without extranodal spread, the 1-, 3-, and 5-year overall survival rates were 98.6%, 89.6%, and 80.8%, respectively, compared with 91.0%, 79.6%, and 68.8% for patients with extranodal spread. Recurrence-free survival was significantly worse in patients with extranodal spread (p = 0.01; Figure 1). The corresponding 1-, 3-, and 5-year recurrence-free survival rates were 93.0%, 79.3%, and 75.0% versus 77.1%, 56.8%, and 56.8%, respectively.

**Figure 1.**
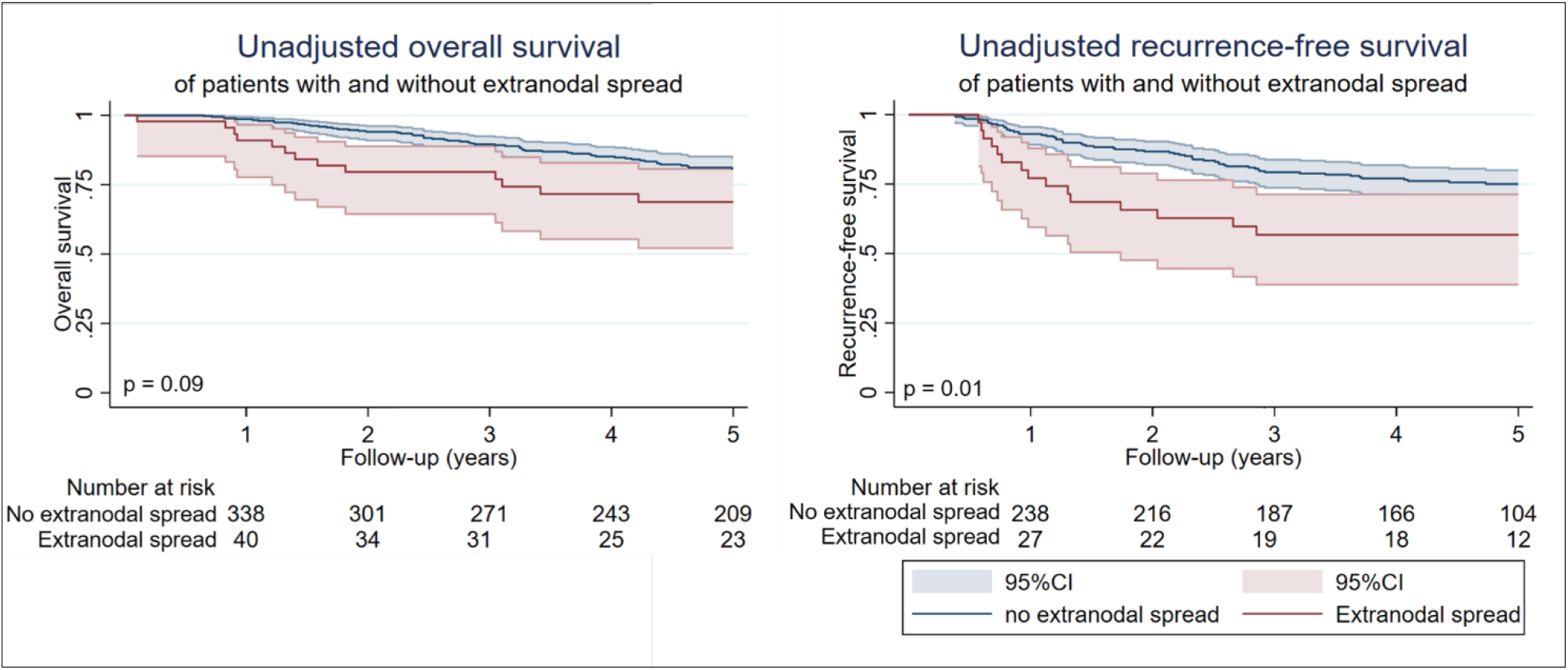
Unadjusted Kaplan-Meier curves for overall survival and recurrence-free survival

After adjusting for age, histology, lympho-vascular space invasion, parametrial invasion and the number of positive lymph nodes, extranodal spread remained significantly associated with recurrence-free survival, but not with overall survival. Patients with extranodal spread had a higher hazard of recurrence compared to those without extranodal spread (HR 1.88; 95%CI: 1.07–3.32). Other variables associated with an increased hazard of recurrence included a non-squamous histological subtype, and a higher number of positive lymph nodes (Table 2).

**Table 2.** Multivariable Cox proportional-hazards analyses.

|  | Overall survival |  |  | Recurrence-free survival |  |  |
| --- | --- | --- | --- | --- | --- | --- |
|  | HR | 95% CI | p-value | HR | 95% CI | p-value |
| Capsular status |  |  |  |  |  |  |
| No extranodal spread | 1.00 |  |  | 1.00 |  |  |
| Extranodal spread | 1.35 | 0.77 - 2.36 | 0.30 | 1.92 | 1.09 - 3.40 | 0.03 |
| Age at time of diagnosis | 1.04 | 1.03 - 1.06 | <0.01 | 1.01 | 0.99 - 1.03 | 0.21 |
| Histology |  |  |  |  |  |  |
| Squamous cell carcinoma | 1.00 |  |  | 1.00 |  |  |
| Adenocarcinoma/Adenosquamous carcinoma | 2.08 | 1.37 - 3.18 | <0.01 | 2.46 | 1.56 - 3.90 | <0.01 |
| Lympho-vascular space invasion |  |  |  |  |  |  |
| Absent | 1.00 |  |  | 1.00 |  |  |
| Present | 1.47 | 0.73 - 2.95 | 0.28 | 1.63 | 0.70 - 3.78 | 0.26 |
| Unknown | 1.37 | 0.49 - 3.83 | 0.54 | 1.23 | 0.36 - 4.26 | 0.74 |
| Parametrial invasion |  |  |  |  |  |  |
| Absent | 1.00 |  |  | 1.00 |  |  |
| Present | 1.17 | 0.69 - 1.96 | 0.56 | 1.47 | 0.82 - 2.62 | 0.18 |
| Number of positive lymph nodes | 1.13 | 1.09 - 1.17 | <0.01 | 1.09 | 1.05 - 1.13 | <0.01 |

HR, hazard ratio; CI, confidence interval.
Overall survival analysis based on patients diagnosed between 2009 and 2024 (n=408).
Recurrence-free survival analysis based on patients diagnosed between 2009 and 2019 (n=307).

In the Bayesian multivariable Cox proportional-hazards model, extranodal spread was associated with worse overall survival, with a HR of 1.63 (95% credible interval 1.14–2.27) and a posterior probability of 99.6% that the HR exceeded 1 (Table 3).

**Table 3.** Bayesian Cox proportional-hazards model with informative prior.

|  | Overall survival |  |  |
| --- | --- | --- | --- |
|  | HR | 95% credibility interval | Posterior probability |
| Capsular status |  |  |  |
| No extranodal spread | 1.00 |  |  |
| Extranodal spread | 1.63 | 1.14 - 2.27 | 0.01 |
| Age at time of diagnosis | 1.04 | 1.03 - 1.06 | <0.01 |
| Histology |  |  |  |
| Squamous cell carcinoma | 1.00 |  |  |
| Adenocarcinoma/Adenosquamous carcinoma | 2.13 | 1.36 - 3.14 | <0.01 |
| Lympho-vascular space invasion |  |  |  |
| Absent | 1.00 |  |  |
| Present | 1.63 | 0.77 - 3.19 | 0.12 |
| Unknown | 1.54 | 0.44 - 3.73 | 0.29 |
| Parametrial invasion |  |  |  |
| Absent | 1.00 |  |  |
| Present | 1.16 | 0.65 - 1.84 | 0.33 |
| Number of positive lymph nodes | 1.13 | 1.08 - 1.17 | <0.01 |

Posterior probabilities from Bayesian analyses should not be interpreted as p-values from frequentist statistics.

**Table 4.** Site of first recurrence within five years after diagnosis.

|  | Total<br>(N=79) |  | No extranodal spread<br>(N=65) |  | Extranodal spread<br>(N=14) |  | p-value |
| --- | --- | --- | --- | --- | --- | --- | --- |
|  | N | (%) | N | (%) |  | (%) |  |
| Locoregional recurrence* | 13 | (16.5%) | 9 | (15.0%) | 4 | (36.4%) | 0.09 |
| Distant recurrence* | 58 | (73.4%) | 51 | (85.0%) | 7 | (63.6%) |  |
| Unknown site | 8 | (10.1%) | 5 |  | 3 |  |  |

\*Patients with simultaneous locoregional and distant recurrence at first recurrence were classified as having distant recurrence.

Among patients who developed a recurrence within five years, 16.5% of patients were classified as having a locoregional recurrence, whereas 73.4% were classified as having distant recurrence. The distribution of recurrence patterns (locoregional vs. distant recurrences) did not significantly differ between patients with and without extranodal spread (p=0.09). Recurrence type was unknown in 5 out of 65 patients without extranodal spread (7.6%) and in 3 out of 14 patients with extranodal spread (21.4%).

The sensitivity analysis showed no differences in survival between patients with assumed absence (i.e. unreported status) of extranodal spread and those with pathologically confirmed absence of extranodal spread (appendix A.2).

## Discussion

### Summary of Main Results

This nationwide, population-based study evaluated the prognostic significance of extranodal spread in patients with clinically early-stage cervical cancer treated with radical hysterectomy and lymph node assessment. Extranodal spread was associated with poorer recurrence-free survival across analyses. For overall survival, the findings were directionally consistent, suggesting an adverse association with extranodal spread. Taken together, these findings suggest that extranodal spread may be an unfavorable prognostic factor in early-stage cervical cancer. No significant differences were found in site (locoregional or distant) of first recurrence.

### Results in the Context of Published Literature

The findings are consistent with observations in other tumor types, where extranodal spread has been linked to more aggressive recurrence patterns. In vulvar cancer, for example, extranodal spread is repeatedly associated with higher rates of distant metastases and reduced locoregional control,^9, 22^ and previous studies in head-and-neck cancer suggest a similar trend.^23-26^ Although tumor biology differs across cancer types, our results align with this broader pattern, supporting the notion that extranodal spread may also carry prognostic relevance in cervical cancer. However, unlike in head-and-neck cancer or vulvar carcinoma, where the presence of extranodal spread informs staging and may prompt consideration of adjuvant (chemo)radiotherapy,^9, 27, 28^ the cervical cancer guidelines do suggest reporting of extranodal spread, but do not address whether and how extranodal spread should guide decisions regarding adjuvant treatment or follow-up.^12, 29^

The limited clinical impact in cervical cancer may reflect inconsistent evidence regarding extranodal spread as an independent prognostic factor, as prior studies report mixed results and are often limited by small sample sizes or highly selected populations.^4, 12^-^15, 30^ The large study by Horn et al., however, demonstrated higher recurrence rates and worse overall survival in patients with extranodal spread, based on a cohort of 256 patients, of whom 79 had extranodal spread. Our analysis aligns with these findings, suggesting that extranodal spread identifies a subgroup at increased risk of recurrence. This may be clinically relevant for risk stratification and for increasing clinical awareness during follow-up and patient counseling. At the same time, recurrence risk in cervical cancer is known to be multifactorial, shaped by variables such as lympho-vascular space invasion, tumor size and depth of invasion,^31, 32^ indicating that extranodal spread contributes prognostic information, but should not be interpreted in isolation. While recommendations for intensified adjuvant treatment remain premature, these results collectively call for renewed discussion within the field on how extranodal spread should inform follow-up planning and patient counseling.

While extranodal spread was negatively associated with recurrence-free survival, no statistically significant association with overall survival was observed in the frequentist analyses. Evidence from previous literature regarding the impact of extranodal spread on overall survival remains inconsistent. Some studies report worse overall survival in patients with extranodal spread,^4, 14, 15^ while others did not find an effect.^12, 13^ In our study, patients with extranodal spread consistently demonstrated lower overall survival over time.^12, 13^ The lack of statistical significance in the frequentist analyses may be explained by limited statistical power due to the small number of events. Nevertheless, the observed effect estimates were consistent in direction and magnitude with those obtained in the Bayesian analysis. When prior evidence from Horn et al.^4^, was incorporated, a high posterior probability of poorer overall survival associated with extranodal spread was observed, supporting an adverse prognostic effect of extranodal spread on overall survival. Therefore, the estimated effect sizes from the frequentist and Bayesian analysis were similar in direction and magnitude, collectively supporting the interpretation that extranodal spread is likely associated with worse overall survival.

In our study, extranodal spread was observed in 11% of patients, substantially lower than the 30–50% reported previously.^4, 13, 15^ This discrepancy may partly reflect patient selection. For instance, one study included a high proportion (>50%) of patients with pT2 disease.^4^ Given that parametrial involvement is associated with an increased risk of extranodal spread, a higher proportion of pT2b tumors may have contributed to the higher prevalence reported in that study. This interpretation is supported by our own data, in which parametrial involvement was strongly associated with extranodal spread. In addition, one study included patients with para-aortic involvement, further inflating reported prevalence.^15^ Previous single-center studies may also have overestimated extranodal spread due to more selective, high-risk populations and more detailed pathological assessment.^4, 13, 15^ In contrast, our nationwide cohort reflects routine clinical practice across multiple centers. Although subtle extranodal spread may be underrecognized, it likely offers a more representative estimate of its prevalence in daily practice.

This nationwide population-based study provides data from real-world clinical practice and allows correction for several confounders. However, inherent to the retrospective observational design of this study, several limitations should be acknowledged. First, capsular status was not explicitly reported in 20.1% of pathology reports. In these cases, absence of reported capsular involvement was assumed to indicate no evidence of extranodal spread. Sensitivity analyses showed no meaningful differences between patients with missing capsular status and those without extranodal spread, supporting the robustness of this assumption. Second, the retrospective design precluded standardized pathological processing and reporting across centers, which may have resulted in underreporting of extranodal spread, particularly given the known challenges in its pathological assessment. However, the nationwide scope of the cohort reflects real-world clinical practice and aligns with the study’s objective to evaluate the prognostic relevance of extranodal spread in routine care. Third, recurrence-free survival was calculated from the date of radical hysterectomy, regardless of subsequent adjuvant treatment. A uniform starting point was chosen to avoid immortal-time bias due to variability in adjuvant treatment duration. Finally, despite the nationwide design, the number of patients with extranodal spread remained limited, restricting the statistical power of frequentist analyses to detect associations with overall survival. To address this limitation and incorporate prior evidence, a Bayesian model was applied, enabling a more informative probabilistic assessment of the association between extranodal spread and overall survival.

### Implications for Practice and Future Research

The present findings suggest that the presence of extranodal spread identifies a subgroup of patients with clinically early-stage cervical cancer at increased risk of recurrence. While routine intensification of follow-up schedules or adjuvant treatment based solely on this study is not currently supported, awareness of extranodal spread may be valuable for risk stratification and informed patient counselling. From a research perspective, these results highlight the need for prospective multicenter studies with standardized pathological assessment of extranodal spread to better define its independent prognostic value. In addition, future research should explore how extranodal spread interacts with other established risk factors and whether it could be incorporated into multivariable prognostic models or decision frameworks for adjuvant treatment and follow-up planning.

## Conclusion

In clinically early-stage cervical cancer, extranodal spread reporting is recommended by the ESGO guidelines, but this information is rarely integrated into clinical decision-making. This disconnect contrasts with the increasing emphasis on efficiency and personalization in modern healthcare. Our findings indicate a poorer recurrence-free and potentially poorer overall survival following radical hysterectomy and lymph node assessment in patients with extranodal spread. Given that extranodal spread is recommended for reporting in current guidelines and association with recurrence-free survival and potentially overall survival, extranodal spread should be considered a clinically relevant factor to inform follow-up strategies and potentially guide adjuvant treatment decisions.

## Data Availability

Data from IKNL are used under license for this study and therefore not publicly available. (Analytical) code(s) are available upon (reasonable) request in consultation with the corresponding author. Data from PALGA are not publicly available due to PALGA regulations and patient privacy but may be accessed upon reasonable request and approval by PALGA. This research did not receive any specific grant from funding agencies in the public, commercial, or not-for-profit sectors.

## Author contributions

R.G.F.M. van der Ven: Conceptualization, data curation, formal analysis investigation, methodology, project administration, writing – original draft, writing – review and editing;

H.H.B. Wenzel: Conceptualization, data curation, investigation, methodology, supervision, writing – review and editing;

M.A. van der Aa: Conceptualization, investigation, methodology, supervision, writing – review and editing;

C.H. Mom: Conceptualization, investigation, methodology, supervision, writing – review and editing;

N.B. Thuijs: Conceptualization, investigation, methodology, supervision, writing – review and editing;

J. van der Velden: Conceptualization, investigation, methodology, supervision, writing – review and editing.

## References

1. Arbyn M, Weiderpass E, Bruni L, de Sanjosé S, Saraiya M, Ferlay J, et al. Estimates of incidence and mortality of cervical cancer in 2018: a worldwide analysis. The Lancet Global Health. 2020;8(2):e191–e203.

2. Cibula D, Raspollini MR, Planchamp F, Centeno C, Chargari C, Felix A, et al. ESGO/ESTRO/ESP Guidelines for the management of patients with cervical cancer – Update 2023*. International Journal of Gynecological Cancer. 2023;33(5):649–66.

3. Matsuo K, Machida H, Mandelbaum RS, Konishi I, Mikami M. Validation of the 2018 FIGO cervical cancer staging system. Gynecologic Oncology. 2019;152(1):87–93.

4. Horn L-C, Hentschel B, Galle D, Bilek K. Extracapsular extension of pelvic lymph node metastases is of prognostic value in carcinoma of the cervix uteri. Gynecologic Oncology. 2008;108(1):63–7.

5. Huang L, Zheng M, Liu J-H, Xiong Y, Ding H, Tang L, et al. Risk factors and prognosis of IB-IIB cervical carcinoma with common iliac lymph node metastasis. Chinese Journal of Cancer. 2010;29(4):431–5.

6. Olthof EP, Mom CH, Snijders MLH, Wenzel HHB, van der Velden J, van der Aa MA. The prognostic value of the number of positive lymph nodes and the lymph node ratio in early-stage cervical cancer. Acta Obstetricia et Gynecologica Scandinavica. 2022;101(5):550–7.

7. Heublein S, Schulz H, Marmé F, Angele M, Czogalla B, Burges A, et al. Extracapsular Lymph Node Involvement in Ovarian Carcinoma. Cancers. 2019;11(7):924–.

8. Mermod M, Tolstonog G, Simon C, Monnier Y. Extracapsular spread in head and neck squamous cell carcinoma: A systematic review and meta-analysis. Oral Oncology. 2016;62:60–71.

9. Velden JVD, Van Lindert ACM, Lammes FB, Kate FJWT, Sie-Go DMDS, Oosting H, et al. Extracapsular growth of lymph node metastases in squamous cell carcinoma of the vulva. The impact on recurrence and survival. Cancer. 1995;75(12):2885–90.

10. Oonk MHM, Planchamp F, Baldwin P, Mahner S, Mirza MR, Fischerová D, et al. European Society of Gynaecological Oncology Guidelines for the Management of Patients with Vulvar Cancer - Update 2023. International Journal of Gynecological Cancer. 2023;33(7):1023–43.

11. Paleri V, Urbano TG, Mehanna H, Repanos C, Lancaster J, Roques T, et al. Management of neck metastases in head and neck cancer: United Kingdom National Multidisciplinary Guidelines. The Journal of Laryngology & Otology. 2016;130(S2):S161–S9.

12. Hale RJ, Buckley CH, Fox H, Wilcox FL, Tindall VR, Logue JP. The morphology and distribution of lymph node metastases in stage IB/IIA cervical carcinoma: relationship to prognosis. International Journal of Gynecological Cancer. 1991;1(5):233–7.

13. Samlal RAK, van der Velden J, Schilthuis MS, González DG, Ten Kate FJW, Hart AAM, et al. Identification of High-Risk Groups among Node-Positive Patients with Stage IB and IIA Cervical Carcinoma. Gynecologic Oncology. 1997;64(3):463–7.

14. Metindir J, Bilir Dilek G. Evaluation of prognostic significance in extracapsular spread of pelvic lymph node metastasis in patients with cervical cancer. European journal of gynaecological oncology. 2008;29(5):476–8.

15. Morice P, Castaigne D, Pautier P, Rey A, Haie-Meder C, Leblanc M, et al. Interest of Pelvic and Paraaortic Lymphadenectomy in Patients with Stage IB and II Cervical Carcinoma. Gynecologic Oncology. 1999;73(1):106–10.

16. Netherlands Comprehensive Cancer O. Netherlands Cancer Registry (NCR).

17. Casparie M, Tiebosch ATMG, Burger G, Blauwgeers H, van de Pol A, van Krieken JHJM, et al. PALGA, the Nationwide Histo- and Cytopathology Data Network and Archive. A Role for Digital Pathology? Cellular Oncology. 2008;30(4).

18. Fritz A, Percy C, Jack A, Sanmugaratnam K, Soblin L, Parkin DM, et al. International classification of diseases for oncology (ICD-O). 3rd ed. 1st revision2013.

19. International Union Against C. TNM Classification of Malignant Tumours. Chichester 2009.

20. McElreath R. Statistical rethinking: a Bayesian course with examples in R and Stan. 2nd ed. Boca Raton (FL): CRC Press; 2020.

21. Altman DG. Practical statistics for medical research. 2nd ed. Boca Ratlon (FL): CRC press; 1999.

22. Aragona AM, Cuneo NA, Soderini AH, Alcoba EB. An analysis of reported independent prognostic factors for survival in squamous cell carcinoma of the vulva: Is tumor size significance being underrated? Gynecologic Oncology. 2014;132(3):643–8.

23. Becker M, van den Brekel MMW, Maroldi R. ESR Bridges: imaging and treatment of extranodal spread in head and neck cancer—a multidisciplinary view. European Radiology. 2024;35(2):640–2.

24. Henson C, Abou-Foul AK, Yu E, Glastonbury C, Huang SH, King AD, et al. Criteria for the diagnosis of extranodal extension detected on radiological imaging in head and neck cancer: Head and Neck Cancer International Group consensus recommendations. The Lancet Oncology. 2024;25(7):e297–e307.

25. Henson CE, Abou-Foul AK, Morton DJ, McDowell L, Baliga S, Bates J, et al. Diagnostic challenges and prognostic implications of extranodal extension in head and neck cancer: a state of the art review and gap analysis. Frontiers in Oncology. 2023;13.

26. Kimura Y, Sumi M, Sakihama N, Tanaka F, Takahashi H, Nakamura T. MR Imaging Criteria for the Prediction of Extranodal Spread of Metastatic Cancer in the Neck. American Journal of Neuroradiology. 2008;29(7):1355–9.

27. Chakrabarty N, Mahajan A. Radiological extranodal extension in head and neck cancers: current evidence and challenges in imaging detection and prognostic impact. BJR|Open. 2024;7(1).

28. Luchini C, Nottegar A, Solmi M, Sergi G, Manzato E, Capelli P, et al. Prognostic implications of extranodal extension in node-positive squamous cell carcinoma of the vulva: A systematic review and meta-analysis. Surgical Oncology. 2016;25(1):60–5.

29. Raspollini MR, Lax SF, McCluggage WG. The central role of the pathologist in the management of patients with cervical cancer: ESGO/ESTRO/ESP guidelines. Virchows Archiv. 2018;473(1):45–54.

30. Tinga DJ, Timmer PR, Bouma J, Aalders JG. Prognostic significance of single versus multiple lymph node metastases in cervical carcinoma stage IB. Gynecologic Oncology. 1990;39(2):175–80.

31. Wenzel HHB, Schnack TH, Van der Aa MA, Jensen PT, Høgdall CK, Hardie AN, et al. Risk factors for lymph node metastasis in women with FIGO 2018 IA cervical cancer with a horizontal spread of > 7 mm. Eur J Cancer. 2024;212:115063.

32. Olthof EP, van der Aa MA, Adam JA, Stalpers LJA, Wenzel HHB, van der Velden J, et al. The role of lymph nodes in cervical cancer: incidence and identification of lymph node metastases-a literature review. Int J Clin Oncol. 2021;26(9):1600–10.

