## Supplementary figures and images for "The impact of extranodal spread on overall and recurrence-free survival in patients with clinically early-stage cervical cancer, but with tumor positive pelvic lymph nodes after radical hysterectomy"

### appendix A.1

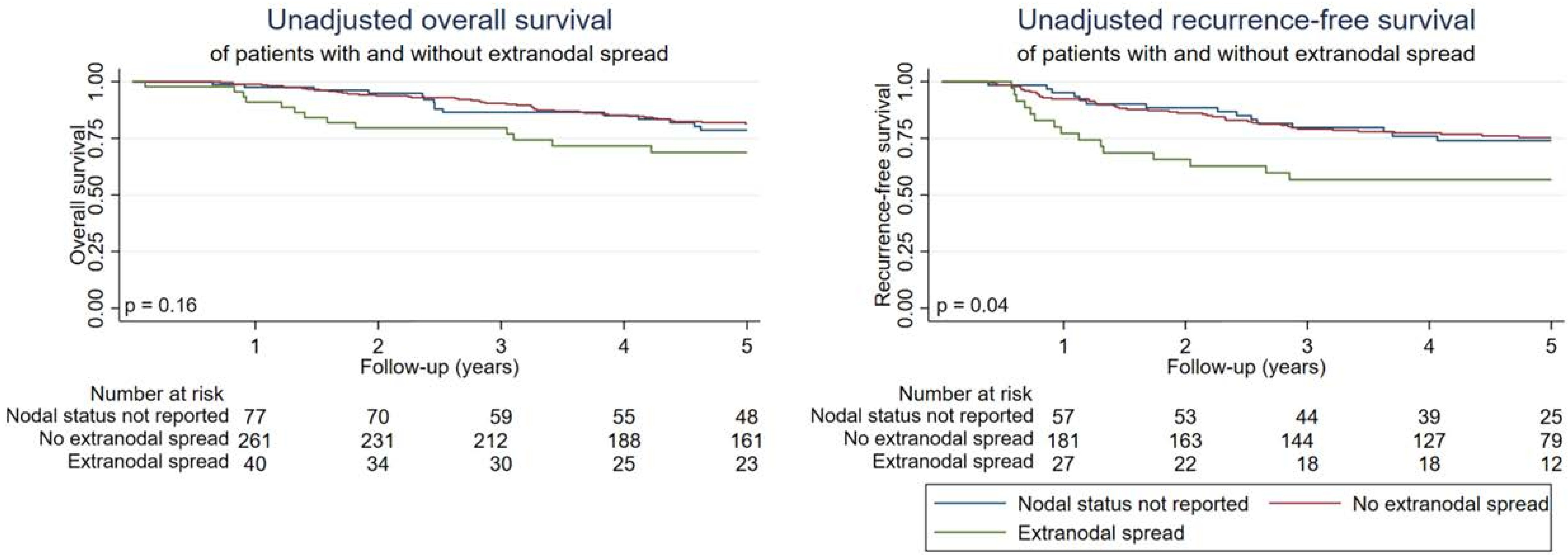
