## appendix A.2 for "The impact of extranodal spread on overall and recurrence-free survival in patients with clinically early-stage cervical cancer, but with tumor positive pelvic lymph nodes after radical hysterectomy"

**Supplementary table 1** Patient- and disease-related characteristics of patients with extranodal spread, without extranodal spread, and with unreported nodal status

| | **Nodal-status not reported** | **No extranodal spread** | **Extranodal spread** | **p-value** |
|---|---|---|---|---|
| N | 82 | 280 | 46 | |
| Year of diagnosis* | 2013 (2010, 2018) | 2016 (2012, 2020) | 2014 (2011, 2018) | <0.01 |
| Age at diagnosis* | 43 (39, 50) | 42 (35, 52) | 43 (38, 55) | 0.61 |
| Tumor stage | | | | |
| 1A1 - 1B3 | 74 (90.2%) | 263 (93.9%) | 42 (91.3%) | 0.43 |
| 2A - 2A2 | 8 (9.8%) | 17 (6.1%) | 4 (8.7%) | |
| Histology | | | | |
| Squamous cell carcinoma | 64 (78.0%) | 191 (68.2%) | 28 (60.9%) | 0.10 |
| Adenocarcinoma / Adenosquamous carcinoma | 18 (22.0%) | 89 (31.8%) | 18 (39.1%) | |
| Differentiation grade | | | | |
| Well-differentiated | 2 (2.4%) | 12 (4.3%) | 3 (6.5%) | 0.82 |
| Moderately differentiated | 31 (37.8%) | 91 (32.5%) | 12 (26.1%) | |
| Poorly differentiated | 25 (30.5%) | 94 (33.6%) | 17 (37.0%) | |
| Unknown | 24 (29.3%) | 83 (29.6%) | 14 (30.4%) | |
| Lympho-vascular space invasion | | | | |
| Absent | 12 (14.6%) | 43 (15.4%) | 2 (4.3%) | 0.13 |
| Present | 63 (76.8%) | 224 (80.0%) | 40 (87.0%) | |
| Unknown | 7 (8.5%) | 13 (4.6%) | 4 (8.7%) | |
| Depth of invasion | | | | |
| <= 3 mm | 5 (6.1%) | 13 (4.6%) | 0 (0.0%) | 0.29 |
| >3 - <=5 mm | 9 (11.0%) | 17 (6.1%) | 5 (10.9%) | |
| >5 mm | 49 (59.8%) | 195 (69.6%) | 33 (71.7%) | |
| unknown / N/A | 19 (23.2%) | 55 (19.6%) | 8 (17.4%) | |
| Parametrial invasion | | | | |
| Absent | 72 (87.8%) | 244 (87.1%) | 33 (71.7%) | 0.01 |
| Present | 9 (11.0%) | 36 (12.9%) | 13 (28.3%) | |
| Unknown | 1 (1.2%) | 0 (0.0%) | 0 (0.0%) | |
| Number of lymph nodes examined* | 22 (18, 32) | 24 (18, 30.5) | 22 (16, 26) | 0.17 |
| Number of positive lymph nodes* | 1 (1, 2) | 1 (1, 2) | 2 (1, 4) | 0.02 |
| Adjuvant therapy | | | | |
| No chemotherapy nor radiotherapy | 5 (6.1%) | 11 (3.9%) | 1 (2.2%) | 0.33 |
| Chemotherapy | 1 (1.2%) | 1 (0.4%) | 0 (0.0%) | |
| Radiotherapy | 25 (30.5%) | 66 (23.6%) | 8 (17.4%) | |
| Chemo- and radiotherapy | 51 (62.2%) | 202 (72.1%) | 37 (80.4%) | |

* Median with interquartile range (IQR)
